# A Brief Burnout Evaluation Scale (BBES) as a potential tool to prevent collapse of the health care task force during the COVID-19 pandemic

**DOI:** 10.1101/2020.09.21.20198804

**Authors:** Tamires Martins Bastos, Gabriela Massaro Carneiro Monteiro, Rogério Boff Borges, Carolina Meira Moser, Daniel Luccas Arenas, Ana Margareth Bassols, Pricilla Braga Laskoski, Simone Hauck

**Affiliations:** Graduation Program in Psychiatry and Behavioral Sciences, Federal University of Rio Grande do Sul, Porto Alegre, RS, Brazil; Hospital de Clínicas de Porto Alegre (HCPA), RS, Brazil; Unidade de Bioestatística, Grupo de Pesquisa e Pós-graduação (GPPG), HCPA, Brasil

**Keywords:** health care professionals, burnout syndrome, COVID-19, depression, suicide

## Abstract

**Introduction:** In the context of the COVID-19 pandemic, where overloaded health systems seem inevitable, there is a need for reliable, conceptually adequate, and easily applied measurement tools to identify health professionals at risk.

**Objective:** to present the preliminary psychometric properties of a Brief Burnout Evaluation Scale (BBES) and its association with important outcomes, i.e., moderate to severe depression and suicidal ideation.

**Methods:** The BBES has 4 Likert-type items and was tested as part of a cross-sectional study that included 401 medical students. Reliability analysis and validity studies were performed.

**Results:** In the parallel analysis, two factors were extracted, explaining 84.4% of the variance. The Cronbach’s alpha was 0.78, showing high internal consistency. Considering a cut-off point of 12, the odds ratio for moderate to severe depression was 3.01 (CI 1.7-5.22; p<0.001) and for last month suicidal ideation 2.96 (CI 1.6-5.48).

**Conclusion:** The results suggest good psychometric characteristics for the BBES, thus reinforcing its utility as an assessment tool for evaluating the well-being or distress of health professionals. It carries with it the potential to implement early interventions and to prevent the descent into burnout so common today in the health care task force during the pandemic.

## Introduction

In the last two decades the health care task force has been recognized as going through an increasing rate of distress and emotional strain. Various combinations of burnout, depression, and suicidal ideation lead to work-related stress as well as other psychiatric symptoms. A recent review showed that exhaustion and mental illness already affect, on average, 30-60% of all physicians and/or physicians-in- training in North America.^1^ These percentages will undoubtedly climb as a result of the additional stressors related to the COVID-19 pandemic, where health professionals face demanding scenarios in which their ongoing capacity to function may be seriously compromised to the point of collapse. A study that evaluated 1257 professionals who treated patients exposed to COVID-19 in China found symptoms of depression in 50.4%, anxiety in 44.6%, insomnia in 34% and distress in 71.5% of the total sample.^2^

In the current context of this pandemic, health care professionals are in desperate need of reliable, conceptually appropriate, and easily applied measurement tools to identify individuals at risk for the Burnout Syndrome. Indeed, burnout as a construct has been hotly debated in the literature.^3^ Some authors assert that it could simply be an alternative to depression that is less stigmatizing to many physicians. However, recent evidence has clarified that burnout and depression are related but distinct constructs. In fact, burnout symptoms may lead to more serious outcomes, including significant depression and suicidal ideation or attempts. Moreover, even when controlling for depression, burnout symptoms result in considerable costs to the individual, the quality of patient care, and to society itself.^1,4-7^ These studies also reveal that the key dimensions that most clearly characterize patients with burnout are emotional exhaustion (EE) and depersonalization (DP) as defined by the Maslach Burnout Inventory (MBI) and/or the Oldenburg Burnout Inventory. Those two dimensions are also the ones more related to depression and psychological distress.^3, 5^ Therefore, an early detection of those symptoms could be used as a marker of risk and perhaps lead to earlier treatment that heads off more severe outcomes. Tools sensitive enough to detect these symptoms can be used to monitor the health professionals, and to implement interventions earlier in the process of the stress continuum. They can also be used to measure the effectiveness of those interventions.

The purpose of this brief communication is to present the preliminary psychometric properties of a Brief Burnout Evaluation Scale (BBES), and its association with significant outcomes, i.e. moderate to severe depression and suicidal ideation. In the current setting, the BBES is designed to function as a tool to identify distress more clearly among health care professionals during the COVID-19 pandemic.

## Methods

The 4 items of BBES were developed from an online survey that evaluated 2537 Brazilian medical students and doctors in October 2019, through the MBI (submitted manuscript). The two EE items (r = 0.87; r = 0.88) and the two DP items (r = 0.81; r = 0.77) of the MBI that were most correlated with the total score of each dimension were adapted and modified to create the BBES items. The BBES is a five-point Likert self-administered scale, ranging from “totally false” (0) to “totally true” (4) (TABLE 1).

**TABLE 1.** Brief Burnout Evaluation Scale (BBES)

ESCALA BREVE DE AVALIAÇÃO DE BURNOUT – versão validada em português Brasileiro
|  | TOTALMENTE<br>FALSO | PARCIALMENTE<br>FALSO | NEM FALSO<br>NEM<br>VERDADEIRO | PARCIALMENTE<br>VERDADEIRO | TOTALMENTE<br>VERDADEIRO |
| --- | --- | --- | --- | --- | --- |
| *1. Eu me sinto emocionalmente sugado pelo trabalho/faculdade.* | ☐ | ☐ | ☐ | ☐ | ☐ |
| *2. Eu me sinto esgotado por causa do trabalho/faculdade.* | ☐ | ☐ | ☐ | ☐ | ☐ |
| *3. Eu me preocupo que o meu trabalho/faculdade esteja me endurecendo emocionalmente.* | ☐ | ☐ | ☐ | ☐ | ☐ |
| *4. Eu fiquei mais insensível em relação às pessoas desde que eu comecei nesse trabalho/faculdade.* | ☐ | ☐ | ☐ | ☐ | ☐ |

|  | TOTALLY<br>FALSE | PARTIALLY<br>FALSE | NEITHER FALSE<br>NOR TRUE | PARTIALLY<br>TRUE | TOTALLY<br>TRUE |
| --- | --- | --- | --- | --- | --- |
| *1. I feel emotionally drained by my work/college.* | ☐ | ☐ | ☐ | ☐ | ☐ |
| *2. I feel exhausted because of my work/college.* | ☐ | ☐ | ☐ | ☐ | ☐ |
| *3. I worry that my work/college is making me emotionally tough* | ☐ | ☐ | ☐ | ☐ | ☐ |
| *4. I became less sensitive to people since I started this work/college.* | ☐ | ☐ | ☐ | ☐ | ☐ |

Totally false = 0; partially false = 1; neither false nor true = 2; partially true = 3; totally true = 4 OBS.: Only the Portuguese version was validated.

In a second step, the BBES was tested as part of a cross-sectional study conducted between November 2019 and March 2020, which included 401 students regularly enrolled in the medical course at the Federal University of Rio Grande do Sul (UFRGS). The data were collected through an online questionnaire on the SurveyMonkey® platform, preserving the anonymity of the participants. The questionnaire contained socio-demographic variables and questions related to mental health (such as suicide ideation in the last month) and the Beck Depression Inventory (BDI).^8^ The studies were approved by the ethics committee of the Hospital de Clínicas de Porto Alegre (CAAE 70231617600005327 and 85311418800005327 respectively).

The Kaiser-Meyer-Olkin index (KMO) and Bartlett’s sphericity test were used to verify whether the data were subject to factor analysis. A polychoric correlation matrix was performed, along with an exploratory factor analysis with oblique rotation to assess the underlying structure of the scale. The number of factors was determined by the method of parallel analyzes. Internal validity was tested using Cronbach’s alpha coefficient.

An initial cut-off point was suggested, considering that, on average, the subject responded “partially true” to the four symptoms. Considering this proposed cut-off point, the odds ratio for moderate to severe depression, according to the BDI^8^, and for suicidal ideation in the last month was estimated using logistic regression model. Further odds tests for moderate to severe depression according to the BDI diagnosis of moderate to severe depression were further performed for cut-off points between 10 and 14 points in the BBES. The analyzes were performed in the R program (r-project.org) using the psych package version 1.9.12. Whenever necessary, a significance level of 5% was adopted.

## Results

The mean age was 23.1 and the standard deviation was 3.3 years. Most of the sample consisted of caucasian students (73.2%) and women (54.9%); 67 students (16.7%) had moderate to severe depression according to the BDI, 48 (12%) suicidal ideation in the last month.

A KMO index 0.64 and a Bartlett sphericity test with p <0.001, showed the adequacy of the use of factor analysis to evaluate the data. In the parallel analysis, two factors were extracted, explaining 84.4% of the total variance. Items EE1 and EE2 presented factorial loads of 0.91 and 0.84 on the first factor, and items DP1 and DP2 loads of 0.75 and 0.80 on the second factor. The estimated correlation between the factors was 0.51. The Cronbach’s alpha for the total score was 0.78, showing high internal consistency.

The score can certainly be used as a continuous variable for several purposes, such as evaluating the staff over a course of time, both to early intervene in the case of health deterioration as well as to evaluate the effectiveness of interventions. However, it is often desirable to establish a cut-off point, at least one to be further tested as valid. Considering a suggested cut-off point of 12 or more, the odds ratio (OR) for moderate to severe depression was 3.01 (CI 1.7-5.22; p<0.001) and for suicidal ideation in the last month 2.96 (CI 1.6-5.48). With a cut-off point of 10 or more the OR for moderate to severe depression was 3.99 (CI 2.11-7.54; p<0.001); with a cut-off point of 11 or more it was 3.73 (CI 2.1-.6.62; p<0.001); with a cut-off point of 13 or more it was 3.94 (CI 2.2-.7.06; p<0.001); and with a cut-off point of 14 or more 5.14 (CI 2.64-10; p<0.001). This finding reinforces what has been shown in the literature - namely, that burnout symptoms can be an important marker of emotional distress. Hence, they can be systematically used to monitor the mental health/well-being of the health care staff.

## Conclusion

This study intends to provide a brief, reliable and free-of-charge scale as soon as possible to assess the presence of symptoms of burnout in students and health professionals in Brazil. This need becomes more urgent in the face of the global crisis caused by the pandemic of COVID-19. Due to the current fragility and overload of different health systems, the early identification of cases is essential, not only to protect professionals from burning out, but also to protect patients from the consequences that it may bring to the quality of care. The clear correlation of the scale with serious outcomes, such as significant depression and suicidal ideation, reinforce the utility of the BBES as an assessment tool for evaluating the continuum from well-being to distress of health professionals, making it possible to implement early interventions and sparing the task force.

The study has limitations, inherent to preliminary studies that need replication in larger samples, other contexts and with more sophisticated methodologies. However, it brings to society, in an urgent moment, a tool with great potential use that can be applied even in remote regions of the country.

## Data Availability

Unpublished data.
The 4 items of BBES were developed from an online survey that evaluated 2537 Brazilian medical students and doctors in October 2019, through the MBI (submitted manuscript).

## References

1. Mihailescu M and Neiterman E.A scoping review of the literature on the current mental health status of physicians and physicians-in-training in North America. BMC Public Health. 2019;19:1363.

2. Lai J, Ma S, Wang Y, Cai Z, Hu J, Wei N et al. Factors Associated With Mental Health Outcomes Among Health Care Workers Exposed to Coronavirus Disease 2019. JAMA Netw Open. 2020;3:e203976.

3. Rotenstein LS, Torre M, Ramos MA, Rosales RC, Guille C, Sen S et al. Prevalence of Burnout Among Physicians: A Systematic Review. JAMA. 2018;320:1131–1150.

4. Mbanga C, Makebe H, Tim D, Fonkou S, Toukam L and Njim T. Burnout as a predictor of depression: a cross-sectional study of the sociodemographic and clinical predictors of depression amongst nurses in Cameroon. BMC Nurs. 2019;18:50.

5. Njim T, Mbanga CM, Tindong M, Fonkou S, Makebe H, Toukam L et al. Burnout as a correlate of depression among medical students in Cameroon: a cross-sectional study. BMJ Open. 2019;9:e027709.

6. Fitzpatrick O, Biesma R, Conroy RM, McGarvey A. Prevalence and relationship between burnout and depression in our future doctors: a cross-sectional study in a cohort of preclinical and clinical medical students in Ireland. BMJ Open. 2019; 9: e023297.

7. Dyrbye LN, Awad KM, Fiscus LC, Sinsky CA, Shanafelt TD. Estimating the Attributable Cost of Physician Burnout in the United States. Ann Intern Med. 2019;170:784–790.

8. Gomes-Oliveira MH, Gorenstein C, Lotufo Neto F, Andrade LH and Wang YP. Validation of the Brazilian Portuguese version of the Beck Depression Inventory-II in a community sample. Braz J Psychiatry. 2012; 34:389–94.

